# Blood-based transcriptomic biomarkers are predictive of neurodegeneration rather than Alzheimer’s disease

**DOI:** 10.1101/2023.09.15.23295651

**Authors:** Artur Shvetcov, Shannon Thomson, Jessica Spathos, Ann-Na Cho, Heather M. Wilkins, Shea J. Andrews, Fabien Delerue, Timothy A. Couttas, Jasmeen Kaur Issar, Finula Isik, Simran Kaur, Eleanor Drummond, Carol Dobson-Stone, Shantel L. Duffy, Natasha M. Rogers, Daniel Catchpoole, Wendy A. Gold, Russell H. Swerdlow, David A. Brown, Caitlin A. Finney

## Abstract

Alzheimer’s disease (AD) is a growing global health crisis, affecting millions and incurring substantial economic costs. However, clinical diagnosis remains challenging, with misdiagnoses and underdiagnoses prevalent. There is an increased focus on putative, blood-based biomarkers that may be useful for the diagnosis, as well as early detection, of AD. In the present study, we used an unbiased combination of machine learning and functional network analyses to identify blood gene biomarker candidates in AD. Using supervised machine learning, we also determine whether these candidates were indeed unique to AD or whether they were indicative of other neurodegenerative diseases Parkinson’s disease (PD) and amyotrophic lateral sclerosis (ALS). Our analyses showed that genes involved in spliceosome assembly, RNA binding, transcription, protein synthesis, mitoribosomes, and NADH dehydrogenase were the best performing genes for identifying AD patients relative to cognitively healthy controls. This transcriptomic signature, however, was not unique to AD and subsequent machine learning showed that this signature could also predict PD and ALS relative to controls without neurodegenerative disease. Combined, our results suggest that mRNA from whole blood can indeed be used to screen for patients with neurodegeneration but may be less effective at diagnosing the specific neurodegenerative disease.

## 1. Introduction

Alzheimer’s disease (AD), the most common form of dementia, is a rapidly growing global medical crisis. With 50 million people currently affected, and at least an additional 10 million cases a year predicted, the global cost to the economy is estimated to be $1.3 trillion USD annually [1]. The clinical diagnosis of AD has remained challenging: 25-30% of patients are misdiagnosed with AD and a further 50-70% of patients with symptoms of AD don’t receive a probable AD diagnosis from their primary care provider [2]. To combat this, there has been an increased focus on the identification of putative, blood-based, biomarkers to aid in the early diagnosis and detection of AD. Diagnostic tests are especially important for identifying patients with AD who may be suitable for clinical trials [3]. The majority of this work has focused on the core pathological hallmarks of AD including amyloid β (Aβ), phosphorylated tau (pTau), and neurofilament light chain (NfL) [4]. Although there has been some demonstrated diagnostic utility towards these pathogenic traits, for example pTau181 [5], there is evidence that these biomarkers increase with age, even in the absence of clinical AD symptoms [4]. Further, available modelling demonstrates the clinical efficacy of these putative biomarkers are performed on a smaller number of patients (often <100), use cohorts with a significant class imbalance (healthy controls > AD), and rarely examine whether their biomarkers are also predictive of other neurodegenerative diseases. This may lead to significant bias and perhaps limit the clinical utility of these models [6, 7]. Therefore, there is an imperative to evaluate the efficacy of additional AD biomarkers outside those traditionally used, using modelling techniques that are more likely to be generalizable.

Advances in high-throughput omics technologies have allowed the measurement of tens of thousands of genes and molecules that are dysregulated in disease states, making them novel tools for identifying biomarkers. However, historically there has been a tendency to use these high-throughput techniques to test *a priori* hypotheses, potentially limiting the full appreciation of their diagnostic capacity. Further, these datasets are often analyzed using arbitrary, yet commonly used fold change and corrected *p*-value thresholds, which can bias the results and their interpretation [8]. One way around these limitations is to distil new perspectives in high-throughput data, without preconceptions, using machine learning. Such an approach represents a unique and effective way to identify novel biomarkers. A small number of prior studies have used this approach to identify novel blood biomarker signatures in AD using transcriptomic and proteomic data [9–15]. Although these studies have shown promising results, there are some limitations that warrant consideration. First, many studies use a very low number of samples (sometimes <20) [10–13, 15]. Small sample sizes in high dimensional data, like patient-derived omics data, can result in significant problems with pattern recognition and model overfitting [16–18]. Furthermore, as mentioned previously, class imbalances where the number of samples in one group significantly outweighs the other, risks biasing the model in a particular direction, often toward the over-represented cohort. In many studies, the number of cognitively healthy controls far exceeds the number of AD patients and is reflected in high specificity (ability to predict healthy control) performance metrics [12]. Additionally, prior investigations utilizing large datasets from public repositories (e.g., Gene Omnibus (GEO) database), do not specify, either in the meta-data or the publication, the diagnostic criteria for AD [13–15, 19], thereby limiting the generalizability of their findings to patients with AD.

There is substantial overlap in genetics, cellular pathways, and even clinicopathological features across neurodegenerative diseases [20–23]. Therefore, an important consideration in biomarker development is whether the signature(s) identified are specific to the disease of interest. Few machine learning studies for AD biomarker development have determined whether the identified signatures are similar or divergent in other neurodegenerative diseases. One study demonstrated that DNA methylome patterns in AD significantly overlap with those of Parkinson’s disease (PD) and amyotrophic lateral sclerosis (ALS) [24]. Another two research groups have reported that certain features (genes) selected by random forest algorithms overlap between neurodegenerative diseases, including AD, PD, ALS, frontotemporal dementia (FTD), Huntington’s disease (HD), and Friedrich’s Ataxia [15, 19]. However, these studies relied on fold change and *p*-value threshold cutoffs and only compared categories of affected transcripts rather than identifying whether their machine learning models, themselves, were able to predict other diseases relative to healthy controls. Thus, it remains unclear if AD predictive biomarker models are useful for diagnosis or whether their signature overlaps other neurodegenerative diseases.

To address this knowledge gap, we analyzed transcriptomic microarray data from whole blood samples of clinically diagnosed AD patients and healthy controls. To move away from fold change and *p*-value-based identification of dysregulated genes, we used an unbiased combination of unsupervised machine learning and functional enrichment analyses to identify these. We then used random forest models to test whether our identified gene biomarker candidates were indeed specific to AD or, using the same models, whether they were generalizable to two other neurodegenerative diseases: PD and ALS.

## 2. Results

### 2.1. Characteristics of the included datasets

Our review of the GEO database identified five whole-blood microarray datasets that met the inclusion criteria: GSE140829, GSE97760, GSE85426, GSE63061, and GSE63060. Two of these, GSE140829 and GSE85426, did not specify the criteria used to diagnose probable AD and were therefore excluded from our analyses. Samples from the included datasets consisted of patients with a diagnosis of possible or probable AD and cognitively normal controls at time of assessment (Table 1). Donor age (ranging from 72-79 years old) and sex were generally well-matched, both across and within datasets (Table 1). Only one of the three datasets specified the ethnicity of their donors: GSE97760 was made up of primarily white donors with two AD donors who were African American [25]. Although all the datasets report measuring RNA integrity (RIN), none reported the cutoff used. Further, none of the datasets reported APOE genotype of the donors.

**Table 1.**
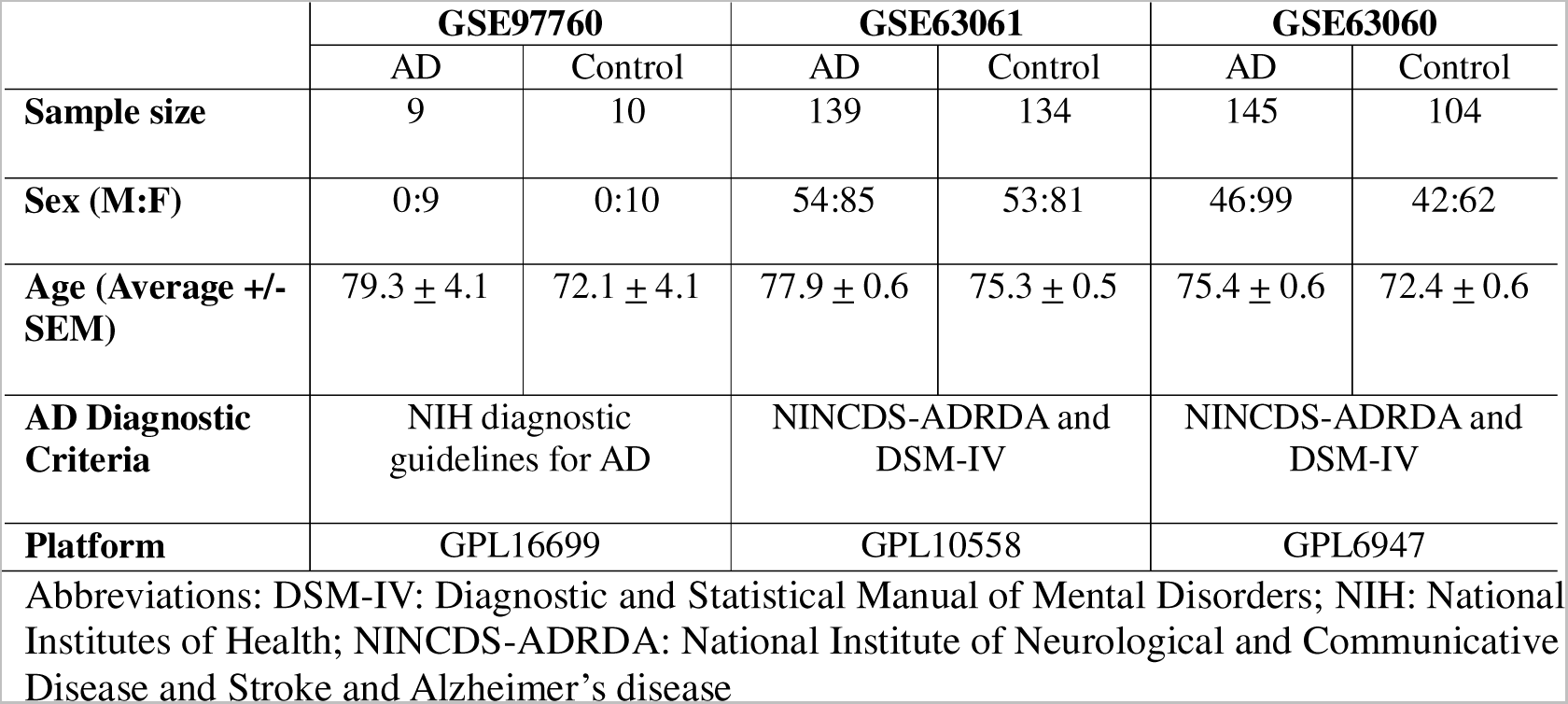
Characteristics of the included AD datasets.

|  | **GSE97760** |  | **GSE63061** |  | **GSE63060** |  |
| --- | --- | --- | --- | --- | --- | --- |
|  | AD | Control | AD | Control | AD | Control |
| **Sample size** | 9 | 10 | 139 | 134 | 145 | 104 |
| **Sex (M:F)** | 0:9 | 0:10 | 54:85 | 53:81 | 46:99 | 42:62 |
| **Age (Average +/- SEM)** | 79.3 ± 4.1 | 72.1 ± 4.1 | 77.9 ± 0.6 | 75.3 ± 0.5 | 75.4 ± 0.6 | 72.4 ± 0.6 |
| **AD Diagnostic Criteria** | NIH diagnostic guidelines for AD |  | NINCDS-ADRDA and DSM-IV |  | NINCDS-ADRDA and DSM-IV |  |
| **Platform** | GPL16699 |  | GPL10558 |  | GPL6947 |  |

Abbreviations: DSM-IV: Diagnostic and Statistical Manual of Mental Disorders; NIH: National Institutes of Health; NINCDS-ADRDA: National Institute of Neurological and Communicative Disease and Stroke and Alzheimer's disease

### 2.2. Principal component and functional network analyses of GSE97760 indicate dysregulated pathways and central gene nodes in whole blood of AD patients

We first performed feature selection using principal component analysis (PCA) on GSE97760 to identify dysregulated central gene nodes that may predict an AD diagnosis. GSE97760 was selected as a reference dataset as all of their AD donors were clinically diagnosed as having advanced AD [25]. PCA showed that there was a distinct separation between the AD and control samples (Figure 1).

**Figure 1.**
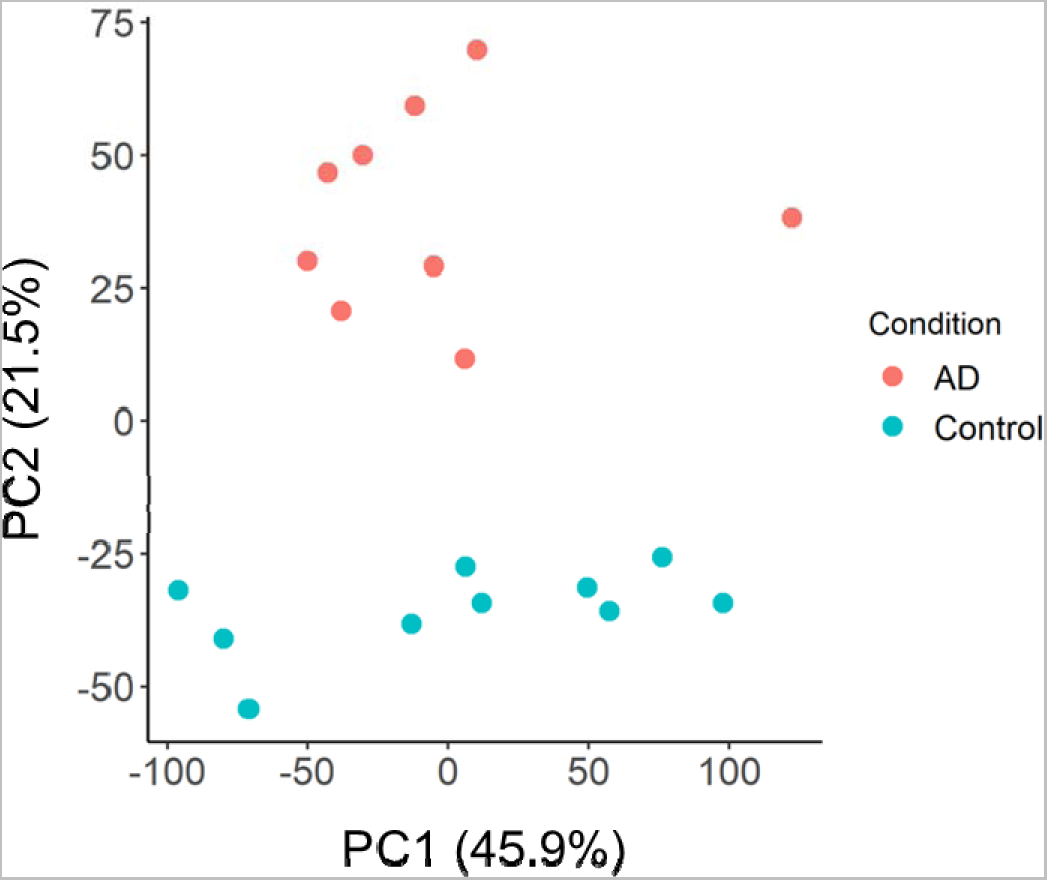
Principal component analysis (PCA) of AD reference dataset GSE97760.

It was also clear that the groups were separating along the y-axis, suggesting that genes within PC2 were playing a key role in driving group separation. Given that PC1, however, contributes to a greater percentage of the variance between groups (45.9% vs 21.5%), we also wanted to ensure that we examined the possibility that genes in PC1 may be good predictors of AD. We therefore took the top 1000 genes that correlated with PC1 (Supplementary Table 1) and PC2 (Supplementary Table 2), respectively, as the most dysregulated genes between AD and controls.

To identify the number of common pathways and interconnections represented in these genes, we performed k-means clustering in STRING [26]. Four clusters were identified as being the optimal number for the dysregulated genes in PC1 and PC2, respectively (Figure 2).

**Figure 2.**
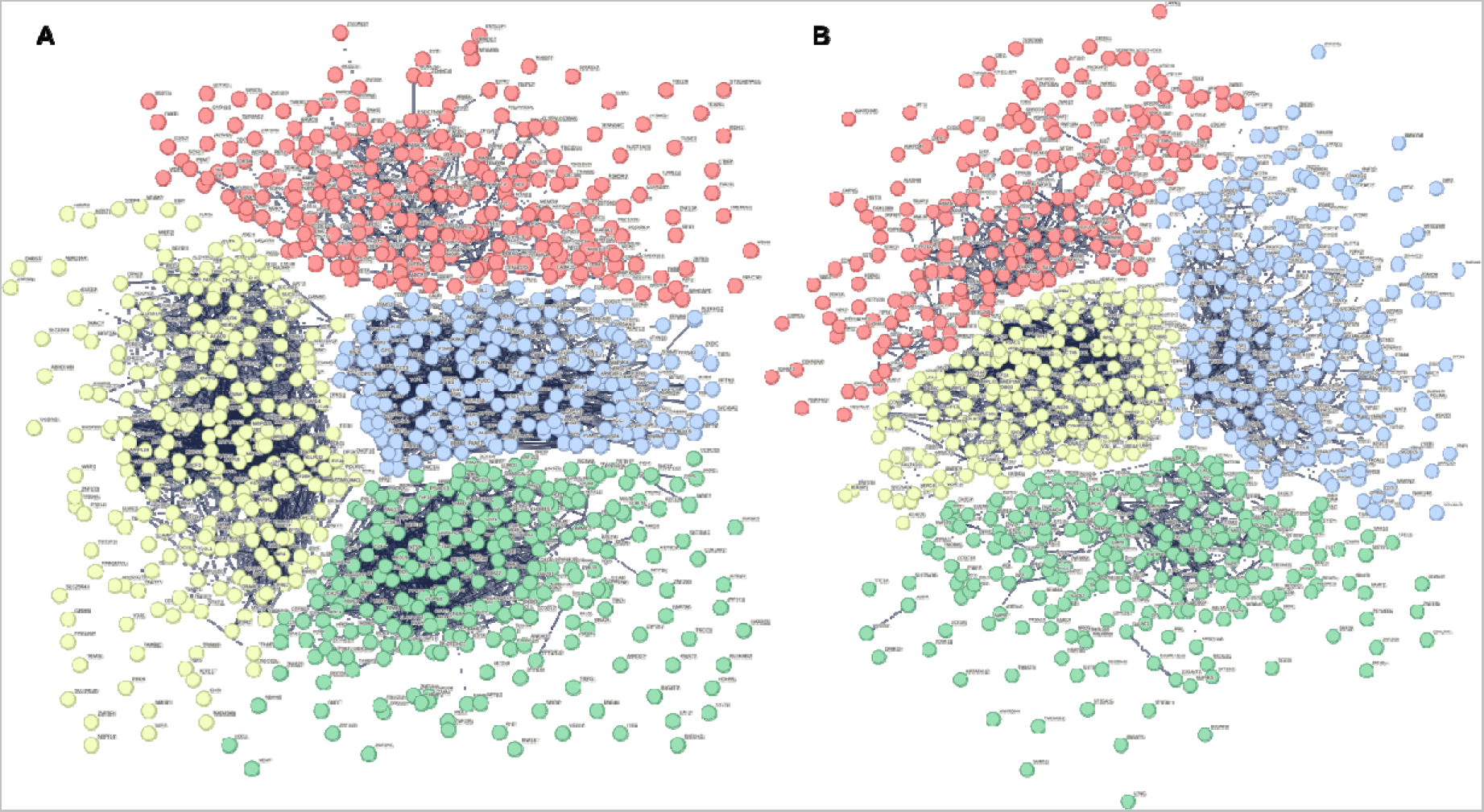
K-means clustering of top 1000 dysregulated genes in GSE97760 identified by principal component analysis. (A) Principal component (PC) 1. Red n = 255, yellow n = 271, green n = 250, and blue n = 224 genes. (B) PC2. Red n = 239, yellow n = 216, green n = 261, and blue n = 284 genes.

Genes within each k-means cluster were then independently examined, again using STRING, to identify overlapping pathways and central gene nodes (the genes that are the most connected within the network) within the cluster. The pathways’ biological function and cellular localization were both characterized using Gene Ontology (GO). In PC1, the first k-means cluster (red) was characterized by genes involved in cellular localization within the endomembrane system (Supplementary Figure 1). There were three central gene nodes identified as playing a role in vesicle formation, the SNARE complex, and signal transduction (Table 2; Supplementary Table 3). The second k-means cluster (yellow) was characterized by 62 central gene nodes involved in metabolic processes across the mitochondrion and ribonucleoprotein complex and included genes from ATP synthase (complex V), mitochondrial ribosomes, and those with roles in mitochondrial respiration (Table 2; Supplementary Table 4; Supplementary Figure 2). The third k-means cluster (green) represented genes important for gene expression in the nucleus (Supplementary Figure 3). The 39 central gene nodes identified here were involved in RNA regulation and transcription (Table 2; Supplementary Table 5). The fourth and final k-means cluster (blue) from PC1 included genes involved in cellular response to stimulus and protein folding, both of which localized to the cytosol (Supplementary Table 6). There were 23 central gene nodes involved in protein folding, maintenance, stabilization, and degradation (Table 2; Supplementary Figure 4).

**Table 2.**
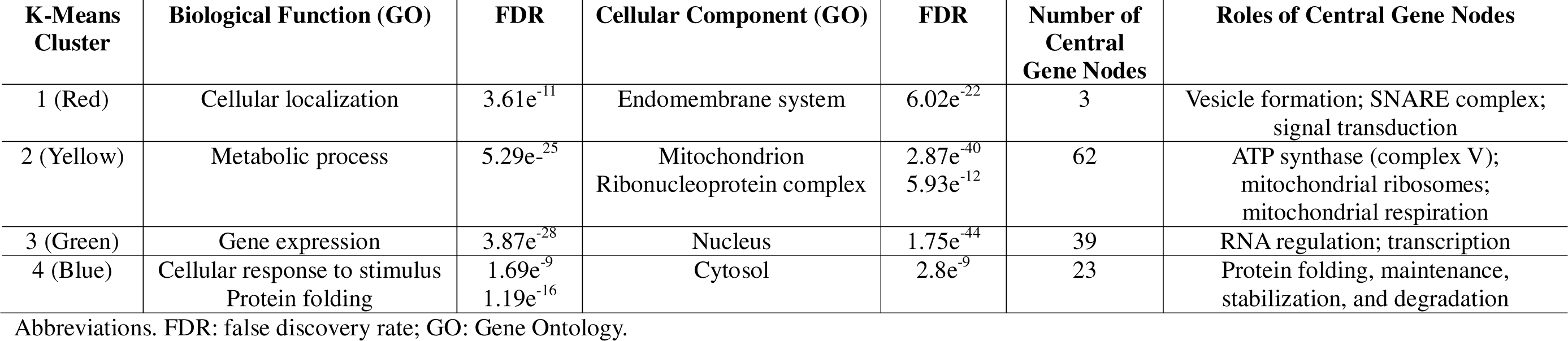
Characterization summary of each k-means cluster from PC1.

We observed a degree of overlap in the biological function of genes between PC1 and PC2 k-means clusters. The first PC2 k-means cluster (red) was similarly characterized by gene expression in the nucleus, however unlike PC1, also included genes expressed in the ribonucleoprotein complex (Supplementary Table 7). Here, the 16 central gene nodes played roles in mRNA regulation and mitochondrial ribosomes (Table 3; Supplementary Figure 5). Genes involved in metabolic processes were also identified in the second PC2 k-means cluster (yellow). These were in both the cytosol and mitochondrion and included 41 central gene nodes involved in cytochrome c oxidase (complex IV), mitochondrial ribosomes, NADH dehydrogenase (complex I), and ribosomes (Table 3; Supplementary Table 8; Supplementary Figure 6). The final two k-means clusters represented pathways unique to PC2. The third k-means cluster (green) included 18 central gene nodes involved in transport within the cytoplasm, including vesicular transport, protein trafficking, and hemoglobin (Table 3; Supplementary Table 9; Supplementary Figure 7). The final k-means cluster (blue) was made up of 12 central gene nodes involved in the regulation of cellular processes in the plasma membrane (Supplementary Figure 8). These included genes with roles in protein kinase signaling, estrogen signaling, transcription, and protein chaperones (Table 3; Supplementary Table 10).

**Table 3.** Characterization summary of each k-means cluster from PC2.

| K-Means Cluster | Biological Function (GO) | FDR | Cellular Component (GO) | FDR | Number of Central Gene Nodes | Roles of Central Gene Nodes |
| --- | --- | --- | --- | --- | --- | --- |
| 1 (Red) | Gene expression | $2.32e^{-9}$ | Nucleus<br>Ribonucleoprotein complex | $1.83e^{-21}$<br>0.00024 | 16 | mRNA regulation; mitochondrial ribosomes |
| 2 (Yellow) | Metabolic process | $2.58e^{-22}$ | Cytosol<br>Mitochondrion | $1.78e^{-16}$<br>$4.40e^{-6}$ | 41 | Cytochrome c oxidase (complex IV);<br>mitochondrial ribosomes; NADH<br>dehydrogenase (complex I); ribosomes |
| 3 (Green) | Transport | $4.58e^{-6}$ | Cytoplasm | $4.19e^{-10}$ | 18 | Vesicular transport; protein trafficking;<br>hemoglobin |
| 4 (Blue) | Regulation of cellular processes | $2.49e^{-10}$ | Plasma membrane | 0.00062 | 12 | Protein kinase signaling; estrogen<br>signaling; transcription; protein<br>chaperone |

Abbreviations. FDR: false discovery rate; GO: Gene Ontology.

### 2.3. Supervised machine learning identifies dysregulated pathways that can predict AD

We used supervised machine learning (random forest) to determine which clusters were the best predictors of AD. Importantly, this was done separately for GSE63061 (dataset A) and GSE63060 (dataset B) to assess the reproducibility and generalizability of our models’ performance. For PC1, the top performing cluster was gene expression, demonstrating a consistently higher sensitivity and precision, average of 0.67 and 0.77, respectively (Table 4; Figure 3).

**Figure 3.**
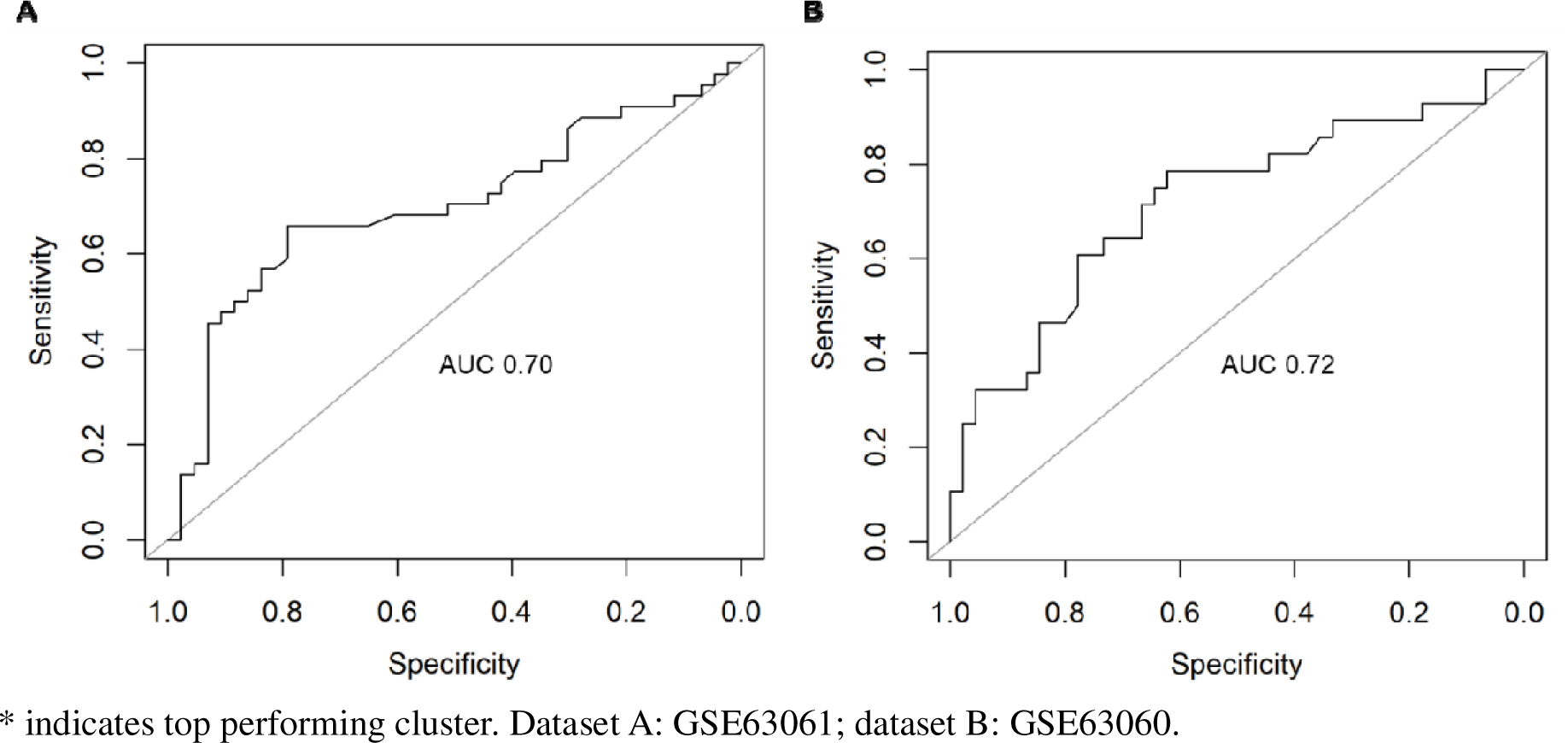
Receiver operating characteristic (ROC) curve of the random forest models’ performance for PC1 gene expression cluster in (**A**) Dataset A, GSE63061 and (**B**) Dataset B, GSE63060. AUC: Area under the curve.

**Table 4.** Random forest performance metrics for the dysregulated clusters from AD PC1.

| Cluster | AUC |  | Sensitivity |  | Specificity |  | Precision |  |
| --- | --- | --- | --- | --- | --- | --- | --- | --- |
|  | A | B | A | B | A | B | A | B |
| Cellular Localization | 0.53 | 0.54 | 0.48 | 0.64 | 0.39 | 0.26 | 0.31 | 0.67 |
| Metabolic Process | 0.55 | 0.79 | 0.54 | 0.72 | 0.46 | 0.81 | 0.51 | 0.89 |
| Gene Expression* | 0.70 | 0.72 | 0.65 | 0.69 | 0.66 | 0.61 | 0.65 | 0.88 |
| Cellular response to stimulus / protein folding | 0.64 | 0.74 | 0.58 | 0.68 | 0.59 | 0.69 | 0.55 | 0.89 |

\* indicates top performing cluster. Dataset A: GSE63061; dataset B: GSE63060.

For PC2, the top performing cluster was metabolic process, with an average sensitivity of 0.7 and precision of 0.77 (Table 5; Figure 4).

**Figure 4.**
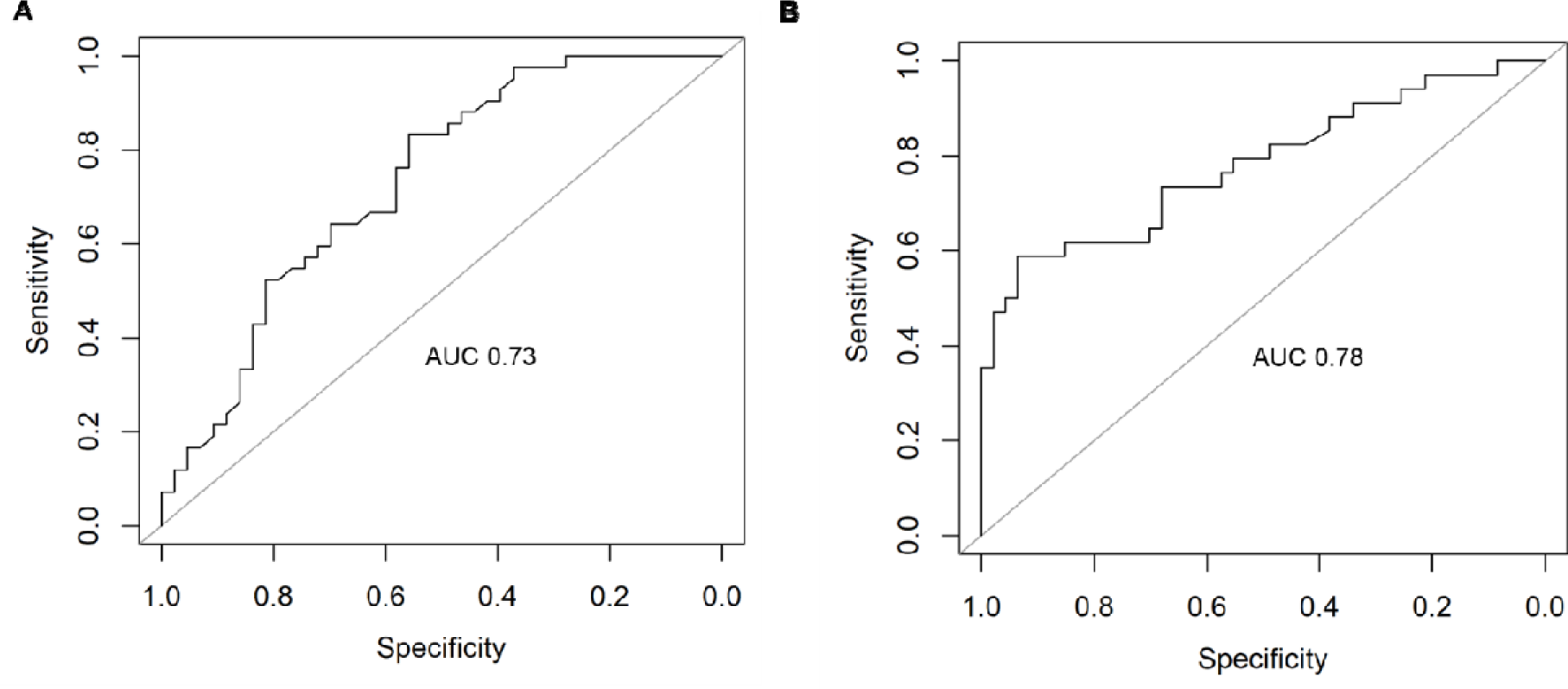
Receiver operating characteristic (ROC) curve of the random forest models’ performance for PC2 metabolic process cluster in (**A**) Dataset A, GSE63061 and (**B**) Dataset B, GSE63060. AUC: Area under the curve.

**Table 5.** Random forest performance metrics for the dysregulated clusters from AD PC2.

| Cluster | AUC |  | Sensitivity |  | Specificity |  | Precision |  |
| --- | --- | --- | --- | --- | --- | --- | --- | --- |
|  | A | B | A | B | A | B | A | B |
| Gene expression | 0.68 | 0.64 | 0.73 | 0.65 | 0.53 | 0.58 | 0.56 | 0.78 |
| Metabolic Process* | 0.73 | 0.78 | 0.65 | 0.75 | 0.67 | 0.72 | 0.7 | 0.83 |
| Transport | 0.58 | 0.43 | 0.55 | 0.60 | 0.48 | 0.55 | 0.55 | 0.87 |
| Regulation of cellular process | 0.57 | 0.58 | 0.54 | 0.67 | 0.42 | 0.41 | 0.40 | 0.64 |

\* indicates top performing cluster. Dataset A: GSE63061; dataset B: GSE63060.

### 2.4. Feature selection of the top performing genes that contribute to AD prediction within each PC cluster

We next sought to identify the top performing gene within each PC cluster that contributed to the random forests’ ability to predict AD. A recursive feature elimination (RFE) was performed on PC1 gene expression (Figure 5A) and PC2 metabolic process cluster (Figure 5B), respectively. At the models’ top performance (blue dot, Figures 5A and 5B), one gene from each cluster dominated the predictive power: LSM3 (PC1 gene expression; Figure 5C), a component of the U4/U6-U5 tri-snRNP complex involved in pre-mRNA splicing and spliceosome assembly, and RPS27A (PC2 metabolic process; Figure 5D), a component of the 40S subunit of the ribosome that plays a role in protein synthesis. The top eight performing genes for the PC1 gene expression cluster included those involved in spliceosome assembly, RNA binding, and transcription (Table 6). On the other hand, the top performers for the PC2 metabolic process cluster included genes involved in protein synthesis, mitoribosomes, and NADH dehydrogenase (Table 6).

**Figure 5.**
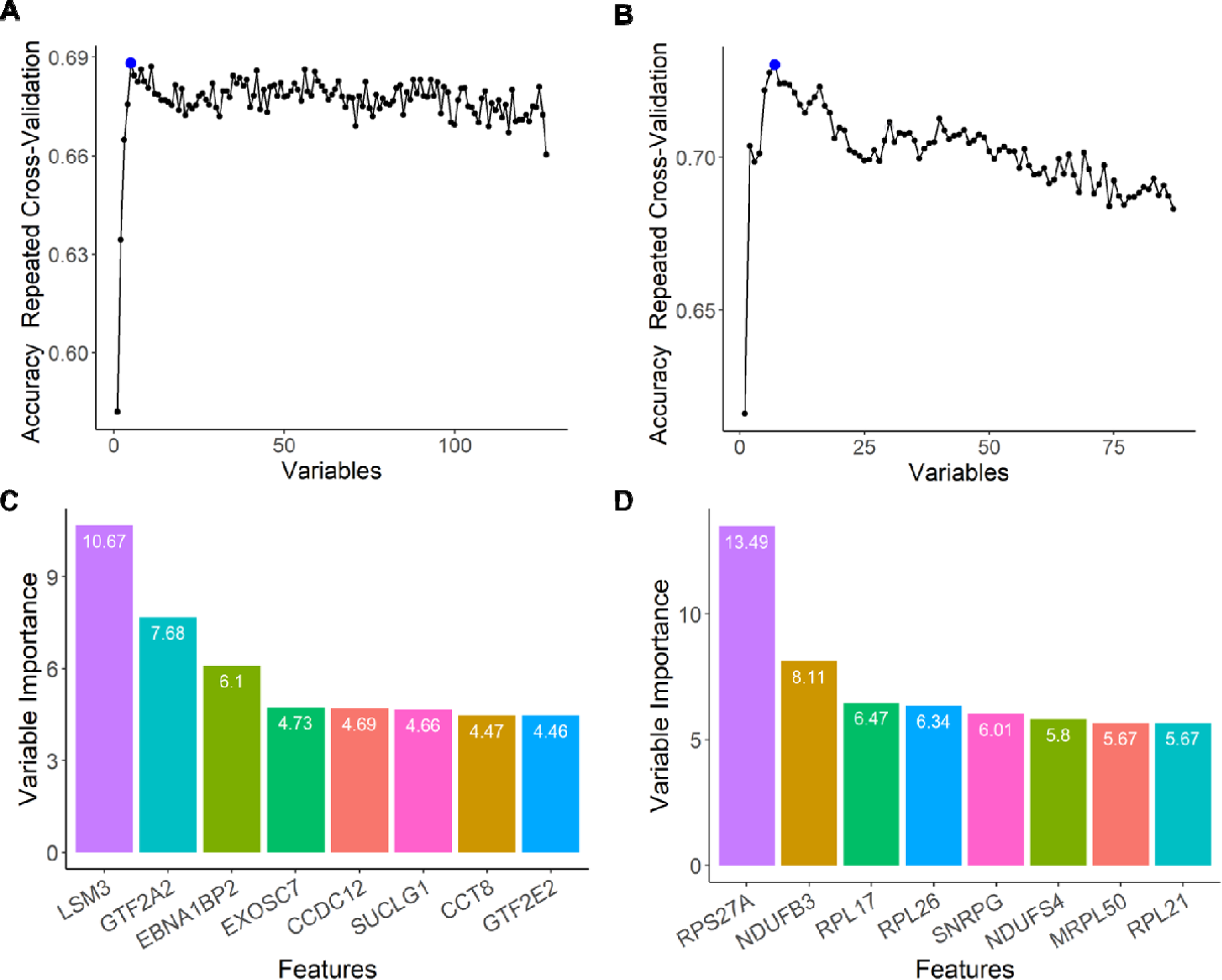
Feature selection of the top performing central gene nodes using recursive feature elimination (RFE) for (**A**) PC1, gene expression cluster and (**B**) PC2, metabolic process cluster. The blue dot indicates peak performance of the model where features were identified from. (**C, D**) The top eight performing genes for the (**C**) PC1 gene expression cluster and (**D**) PC2 metabolic process cluster.

**Table 6.** Biological function of the top performing genes for PC1 gene expression cluster and PC2 metabolic process cluster.

| Abbreviation | Name | Function (NCBI) |
| --- | --- | --- |
| **PC1 Gene Expression Cluster** |  |  |
| LSM3 | U6 snRNA and mRNA Degradation Associated | Component of the U4/U6-U5 tri-snRNP complex involved in pre-mRNA splicing and spliceosome assembly |
| GTF2A2 | General transcription factor IIA subunit 2 | Component of RNA polymerase II transcription machinery, role in transcriptional activation |
| EBNA1BP2 | EBNA1 binding protein 2 | RNA binding activity |
| EXOSC7 | Exosome component 7 | Enables RNA binding and 3'-5'-exoribonuclease activity and RNA metabolism |
| CCDC12 | Coiled-coil domain containing 12 | Component of U2-type and post-mRNA release spliceosomal complexes |
| SUCLG1 | Succinate-CoA ligase<br>GDP/ADP-forming subunit<br>alpha | Alpha subunit of the heterodimeric enzyme succinate coenzyme A ligase, catalyzes conversion of succinyl CoA and ADP to succinate and ATP or GDP to GTP |
| CCT8 | Chaperone containing TCP1<br>subunit 8 | Component of molecular chaperonin-containing T-complex (TRiC), assists in folding of proteins upon ATP hydrolysis |
| GTF2E2 | General transcription factor IIE<br>subunit 2 | Component of RNA polymerase II transcription initiation complex, involved in recruitment of general transcription factor IIH to initiation complex and stimulation of RNA polymerase II C-terminal domain kinase and DNA-dependent ATPase activity |
| **PC2 Metabolic Process Cluster** |  |  |
| RPS27A | Ribosomal protein 27A | Component of 40S subunit of the ribosome involved in protein synthesis |
| NDUFB3 | NADH:ubiquinone<br>oxidoreductase subunit B3 | Component of accessory subunit of mitochondrial membrane respiratory chain NADH dehydrogenase (complex I) |
| RPL17 | Ribosomal protein L17 | Component of the ribosomal 60S subunit, involved in protein synthesis |
| RPL26 | Ribosomal protein L26 | Component of the ribosomal 60S subunit, involved in protein synthesis |
| SNRPG | Small nuclear ribonucleoprotein<br>polypeptide G | Component of the U1, U2, U4, and U5 small nuclear ribonucleoprotein complexes, involved in processing of 3' end of histone transcripts |
| NDUFS4 | NADH:ubiquinone<br>oxidoreductase subunit S4 | Component of nuclear-encoded accessory subunit of mitochondrial membrane respiratory chain NADH dehydrogenase (complex I) |
| MRPL50 | Mitochondrial ribosomal protein<br>L50 | Encodes a putative 39S subunit protein of mitochondrial ribosomes (mitoribosomes) |
| RPL21 | Ribosomal protein L21 | Component of the ribosomal 60S subunit, involved in protein synthesis |

### 2.5. Genes that predict AD are also predictive of neurodegenerative diseases

When identifying potential new biomarkers, it is important to determine if they are disease specific (i.e., AD). We therefore sought to test whether our top-performing gene candidates for AD were also predictive of Parkinson’s disease (PD) and amyotrophic lateral sclerosis (ALS), among the most common neurodegenerative diseases. More specifically, we wanted to determine whether our top performing genes were also able to predict PD and ALS patients irrespective of whether they were trained using an AD dataset (AD train and PD or ALS test) or disease-specific dataset (PD or ALS train and PD or ALS test). For Parkinson’s disease, we sourced two microarray datasets from GEO, GSE6613 (PD vs. healthy control) and GSE72267 (drug-naïve PD vs. healthy control) (Table 7). We also identified one ALS microarray dataset, GSE112681 (ALS vs. healthy control) (Table 7).

**Table 7.** Characteristics of the PD and ALS datasets.

|  | **GSE6613** |  | **GSE72267** |  | **GSE112681** |  |
| --- | --- | --- | --- | --- | --- | --- |
|  | PD | Control | PD | Control | ALS | Control |
| **Sample size** | 50 | 22 | 40 | 20 | 167 | 137 |
| **Sex (M:F)** | 39:11 | 11:11 | 22:18 | 10:10 | 96:68 | 79:58 |
| **Age (Average +/- SEM)** | 69.4 ± 1.2 | 64.4 ± 2.3 | 68.8 ± 1.1 | 60.3 ± 1.3 | Not reported | Not reported |
| **Diagnostic Criteria** | United Kingdom Parkinson's Disease Society Brain Bank clinical diagnostic criteria |  | United Kingdom Parkinson's Disease Society Brain Bank clinical diagnostic criteria |  | Revised El Escorial criteria |  |
| **RNA Quality (RIN average +/- SEM)** | Not specified |  | RIN ≥ 8 |  | RIN ≥ 7 |  |

Random forest models for the top AD gene performers from the PC1 gene expression cluster and PC2 metabolic process cluster, respectively, were trained using a collapsed AD dataset made up of samples from GSE63061 and GSE63060. The trained models were then tested on each of the three PD and ALS datasets, GSE6613, GSE72267, and GSE112681. The random forest models for the PC1 gene expression cluster had precision metrics of >0.65, suggesting that they were able to identify true PD and ALS cases (Table 8).

**Table 8.** Random forest PC1 gene expression cluster performance metrics for neurodegenerative diseases (trained on AD and tested on PD or ALS).

| **Dataset** | **AUC** | **Sensitivity** | **Specificity** | **Precision** |
| --- | --- | --- | --- | --- |
| Parkinson's disease (GSE6613) | 0.48 | 0.69 | 0.30 | 0.69 |
| Parkinson's disease, drug naïve (GSE72267) | 0.54 | 0.72 | 0.39 | 0.65 |
| Amyotrophic lateral sclerosis (GSE112681) | 0.50 | 0.55 | 0.47 | 0.73 |

The models for the PC2 metabolic process cluster had precision metrics of >0.68 and went as high as 0.83, similarly demonstrating that these genes were predictive of PD and ALS (Table 9).

**Table 9.** Random forest PC2 metabolic process cluster performance metrics for neurodegenerative diseases (trained on AD and tested on PD or ALS).

| **Dataset** | **AUC** | **Sensitivity** | **Specificity** | **Precision** |
| --- | --- | --- | --- | --- |
| Parkinson's disease (GSE6613) | 0.52 | 0.68 | 0.27 | 0.68 |
| Parkinson's disease, drug naïve (GSE72267) | 0.61 | 0.71 | 0.45 | 0.83 |
| Amyotrophic lateral sclerosis (GSE112681) | 0.52 | 0.63 | 0.15 | 0.72 |

It is worth noting that while all the models for the PC1 gene expression and PC2 metabolic process clusters had good predictive value for the neurodegenerative disease patients (indicated by high sensitivity and precision), they were unable to identify the healthy controls in each dataset (indicated by low specificity and AUC).

To further validate these findings, we tested our models’ performance when both trained and tested within the same disease dataset (70% training, 30% withheld for testing). This improved all performance metrics for both the PC1 gene expression cluster (Table 10) and PC2 metabolic process cluster (Table 11). Furthermore, it also improved the ability of the models to accurately identify healthy controls (higher specificity) (Tables 10 and 11).

**Table 10.**
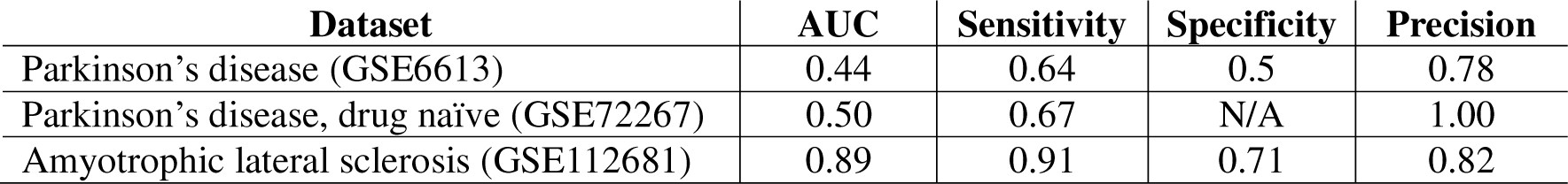
Random forest PC1 gene expression cluster performance metrics for neurodegenerative diseases (trained and tested on same disease).

| Dataset | AUC | Sensitivity | Specificity | Precision |
| --- | --- | --- | --- | --- |
| Parkinson’s disease (GSE6613) | 0.44 | 0.64 | 0.5 | 0.78 |
| Parkinson’s disease, drug naïve (GSE72267) | 0.50 | 0.67 | N/A | 1.00 |
| Amyotrophic lateral sclerosis (GSE112681) | 0.89 | 0.91 | 0.71 | 0.82 |

**Table 11.** Random forest PC2 metabolic process cluster performance metrics for neurodegenerative diseases (trained and tested on same disease).

| Dataset | AUC | Sensitivity | Specificity | Precision |
| --- | --- | --- | --- | --- |
| Parkinson’s disease (GSE6613) | 0.86 | 0.86 | 0.67 | 0.80 |
| Parkinson’s disease, drug naïve (GSE72267) | 1.00 | 1.00 | 1.00 | 1.00 |
| Amyotrophic lateral sclerosis (GSE112681) | 0.89 | 0.91 | 0.74 | 0.84 |

## 3. Discussion

An affirmative diagnosis for neurodegenerative remains difficult and elusive. Currently, there is a reliance on neuroimaging and measurements of biomarkers in the cerebrospinal fluid (CSF) [27]. While these do provide clinical utility, there are caveats, namely accessibility to neuroimaging (particularly in lower-socioeconomic countries) and the perceived invasiveness of CSF collection [27, 28]. Further, targeted proteomic or metabolomic methods for analyzing CSF are costly and methodologically challenging [27]. PCR-based approaches to examine mRNA changes are a favorable alternative given that these assays are timely, reliable, robust, relatively simple, and cost-efficient [29]. However, putative biomarker measurements employing this approach are still in their infancy, hence the need to explore the possibility of reliable blood mRNA biomarkers. Here, we examined publicly available microarray data using machine learning to determine if whole blood transcriptomic signatures are unique to AD or whether they are reflective of other neurodegenerative diseases, including PD and ALS. Our results suggest that mRNA from whole blood can indeed be used to screen for patients with neurodegeneration but may be less effective at diagnosing the specific disease.

Our unsupervised machine learning (PCA and k-means clustering) and functional enrichment analyses indicated there are multiple dysregulated pathways and central gene nodes in the blood of AD patients. Dysfunctional metabolic processes in the mitochondria, cytosol, and ribonucleoprotein complex were found across both PCs, highlighting that these processes are likely dysregulated in the periphery of neurodegenerative disease patients. Central gene nodes involved in these processes including those for NADH dehydrogenase (complex I), cytochrome oxidase C (complex IV), and ATP synthase (complex V) as well as those involved in mitoribosomes and mitochondrial respiration. Similarly, gene expression was found to be dysregulated across the nucleus and ribonucleoprotein complex, implicating processes such as RNA regulation, transcription, and mitoribosome function. Unsurprisingly, both the gene expression and metabolic process central gene nodes were found to be the top performing clusters across PC1 and PC2, respectively, showing an acceptable level of sensitivity and precision. Additional feature selection using RFE identified that the sixteen (eight from PC1 gene expression cluster and eight from PC2 metabolic process cluster) central gene nodes drove the models’ predictive performances. These included those involved in spliceosome assembly, RNA binding, transcription, protein synthesis, mitoribosomes, and NADH dehydrogenase. Despite the AD transcriptomic signatures identified, subsequent machine learning demonstrated that these were not unique to AD. Our models using these top performing genes were also able to predict PD and ALS patients irrespective of whether they were trained using an AD dataset (AD train and PD or ALS test) or disease-specific dataset (PD or ALS train and PD or ALS test). Importantly, the medications commonly used to treat the symptoms of AD, PD, and ALS are different and the ability of our models to perform across neurodegenerative diseases suggests that our findings are not simply an artefact of drug treatments. It is worthy to note, however, that only one of the datasets specified drugs used by the donors: the drug-naïve PD dataset [30]. Although our models showed strong metrics, this dataset was small (n=60). Future research, therefore, should include current prescription and non-prescription data for their donors to ensure that these can be controlled for as well as larger numbers to enable more generalizable conclusions.

Many of the genes we found were good predictors of AD, PD, and ALS have also been previously identified as being broadly implicated in neurodegeneration. RPS27A has been linked to mild cognitive impairment (MCI) and AD [31, 32]. It has also been shown to interact with tau and lead to microglial activation that triggers subsequent widespread neurodegeneration [33, 34]. Both MRPL50 and NDUFB3 have been implicated in AD, glaucoma, and age-related neurodegeneration [35, 36]. In addition to being identified as a gene that links MCI progressing to AD [31], LSM3 has been implicated in PD [37], AD [36], the adult-onset neurodegenerative disorder Fragile X, and Tremor and Ataxia syndrome [38]. SUCLG1 is decreased in AD brains [36] and mutations in this gene have been linked to encephalomyopathic mitochondrial DNA (mtDNA) depletion syndromes characterized by hypotonia and pronounced neurological symptoms [39, 40]. Interestingly, mtDNA is well-documented to play a role across neurodegenerative diseases including, but not limited to, AD, PD, and ALS [41, 42], suggesting that SUCLG1 may be an interesting gene to look at further in the context of neurodegeneration. Further, we implicated NDUFS4 in our neurodegeneration predictive models, which has been connected to both AD [36] and in neurodegeneration associated with the mitochondrial disorder Leigh Syndrome [43, 44].

While we provide strong evidence that mRNA signatures can be used to indicate whether neurodegeneration is present, there are some limitations to this work. First, we used a specific feature selection method (unsupervised machine learning and functional enrichment analyses) to narrow down a list of >15,000 genes to only 16. Therefore, it may be the case that we have overlooked other genes and genetic signature(s) with greater sensitivity towards AD or other neurodegenerative disease. However, this may be mitigated by our selection approach. Our PCA identified the genes highly correlated with the principal components – i.e. those with the greatest influence on driving group separation between AD patients and healthy controls. Inherent to this is that the non-identified genes contribute less to this differentiation and are, therefore, highly likely to be poor predictors of AD. Further, our deliberate exclusion of *p*-value and fold change thresholds ensures that our results weren’t biased toward arbitrary cutoffs. The use of functional enrichment analyses to uncover central gene nodes (the genes that are the most connected within the network) should enrich for genes whose dysregulation have the highest disruptive potential for the system or pathway. In doing so, we may also inadvertently examine genes that are functionally connected to these central gene nodes. In this systems biology approach, dysregulation of a central, highly connected gene is also likely to disrupt downstream connections. Despite this, it can be argued that our functional enrichment analysis is biased toward what is already known and that genes with a subtle influence may be unique to neurodegenerative diseases. These subtleties are unlikely to be uncovered using tools like databases and machine learning; future research would therefore benefit from identifying other methods to examine these.

Another potential limitation of this work is that our models generally had a low specificity and high precision, indicating that the models were poor at identifying true negatives (people without neurodegeneration). In the AD datasets, all healthy controls were in their early to mid-70s and may, therefore, demonstrate subclinical, age-related non-pathological neurodegeneration [45]. This conclusion is strengthened by two findings from our data. First, our models’ sensitivity and precision metrics were higher for the AD dataset GSE63060 relative to GSE63061 where the age of controls was 72 as compared to 75, respectively. In the future, research would benefit from testing whether blood-based biomarkers lose sensitivity and precision with increasing patient age. Second, when our models were trained on the AD datasets and then tested on the PD or ALS datasets, whose healthy controls were around 10 years younger, the specificity was very low. When we instead trained and tested the models on the same dataset (PD or ALS, respectively), the specificity improved dramatically. This also suggests that our models are readily able to differentiate between a patient with neurodegeneration and a healthy control who is younger.

It is also important to consider that there are likely to be sex differences in neurodegenerative disease pathogenesis and therefore biomarkers [46]. For example, a recent study demonstrated that plasma phospho-tau threonine 217 (p-Tau217) and NfL levels differed between males and females with autosomal dominant AD [47]. From a diagnostic testing perspective, however, it is important to identify sex-independent biomarkers that can be routinely used in the clinic. In the present study, although we used a female-only AD dataset as our reference to identify dysregulated central gene nodes, our models still showed good performance (sensitivity and precision) when trained and tested using AD, PD, and ALS datasets made up of both males and females. This suggests that although we used females as a reference point, we have identified genes that are less likely to be sex specific and may therefore be clinically useful to identify patients with neurodegeneration.

The final limitation of this work is that the datasets used in the present study are not derived from single-cell RNA-sequencing, which may limit our interpretation of results. Whole blood is made up of many cell types, including red blood cells, white blood cells (lymphocytes, monocytes, and granulocytes), and platelets. Mature red blood cells are enucleated and, therefore, have low levels of mRNA, with estimates that only about 10% contain mRNA [48, 49]. This suggests that the bulk of mRNA comes from the various white blood cells or platelets [50, 51], highlighting that there may be peripheral dysfunction in these diseases driven by these cell types. For example, CD49^+^ Tregs, relative to other immune cell subsets, have been shown to be increased in blood samples of PD patients relative to healthy controls [52]. This suggests that the relative proportion of blood cell types in a sample may influence the identification of disease-related changes in gene expression, potentially limiting clinical utility [51]. This may especially be the case in patients with comorbid conditions requiring immunosuppression, like cancer patients undergoing chemotherapy. A recent study, however, demonstrated that there were marked differences in CD4^+^ memory, CD4^+^ activated, and CD8^+^ naïve cells, as well as CD38^+^CD16^low^ monocytes, between AD and PD patients [53]. Given that our models were reasonable across AD and PD, as well as ALS, the relative abundance of cell subsets may not provide diagnostic utility with respect to neurodegeneration. Additionally, it is important to note that we do not know if our dysregulated genes are from the CNS or only the periphery. Extracellular vesicles (EVs) package mRNA and may cross the blood brain barrier into general circulation. There is a poor understanding, however, of how EVs do this in a bidirectional manner (i.e. cross from brain to blood and vice versa) [54, 55] therefore we can’t conclude from these data the relative proportion of EVs. Future studies would greatly benefit from dissecting this further to determine whether these blood-based genetic changes are driven by immune cell subtypes or whether they are cell subtype-independent.

In summary, we report a machine learning and functional enrichment analysis approach to identifying whole blood transcriptomic signatures in AD. We also investigated whether these transcriptomic signatures were unique to AD or whether they were indicative of neurodegenerative disease, more broadly. The top performing genes for predicting AD included those involved in spliceosome assembly, RNA binding, transcription, protein synthesis, mitoribosomes, and NADH dehydrogenase. Although we did identify a blood-based transcriptomic signature that was predictive of AD, we found that it could also be used to identify PD or ALS patients relative to non-neurodegenerative disease controls. Together, these findings suggest that there is a shared blood molecular signature across neurodegenerative diseases. This highlights that mRNA from whole blood can likely from whole blood can likely be used to screen patients for neurodegeneration but is less effective at diagnosing the specific neurodegenerative disease. Our identified gene signature should be experimentally investigated to determine whether it is indeed a viable clinical screening test to identify neurodegenerative diseases in patients.

## 4. Materials and Methods

### 4.1. Identification of publicly available transcriptomic datasets

We identified publicly available transcriptomic datasets using a systemic search of the Gene Expression Omnibus (GEO) database. The key term used for the search was “Alzheimer’s disease”, and the results were limited to *homo sapiens*. Datasets were included on the basis that they (a) examined gene expression in whole blood samples, (b) used microarray to generate high throughput transcriptomic data, (c) clinically confirmed AD diagnosis, and (d) included cognitively normal healthy controls. We excluded datasets generated by RNA-sequencing due to their small sample sizes, which increases the risk of developing overfitted, ungeneralizable models using our machine learning methods. Three AD datasets were included: GSE97760, GSE63061, and GSE63060.

### 4.2. Data processing

After downloading the identified GEO datasets, we first confirmed the data was pre-normalized and did not need additional normalization. Log transformations were not used as they are not required in the absence of traditional statistical and fold change analyses. The three datasets were processed independently, duplicate gene entries were removed, and a z-score was calculated for each gene. A list of genes identified in each dataset was generated, and genes not common to all three datasets were discarded. Data processing, merging, and PCA were done in R Studio v1.2.5033 (R v3.6.3) using GEOquery, dplyr, PCA and visualization was done using ggplot.

### 4.3. Feature selection using unsupervised machine learning methods and functional enrichment analyses on GSE97760

We first examined the three AD datasets and selected a ground truth dataset, GSE97760. This was selected due to all the patients having a diagnosis of advanced probable AD, thereby reducing the chances of clinical misdiagnosis [25]. Further supporting this, a PCA indicated a clear group separation between the AD cases and healthy controls (Figure 1). Using GSE97760, we then performed our established two-stage machine learning pipeline as previously described in Finney et al. [8]. Briefly, we first analyzed all genes (>15,000) using PCA (Figure 6). Importantly, PCA allows us to reduce dimensionality and minimize potential information loss [56] while simultaneously revealing hidden patterns in high throughput transcriptomic data [57]. PCA was performed in R Studio v1.2.5033 (R v3.6.3) and visualized using ggplot. The top 1000 genes correlating with PC1 and PC2 were identified and selected as potential biomarker gene candidates (Figure 6). The top 1000 genes were then entered into STRING v11 [26, 58] for gene enrichment analysis and to identify interaction networks. We first performed unsupervised machine learning through k-means clustering to identify the presence of overlapping functional network clusters in the genes (Figure 6). Each k-means cluster was then independently entered into STRING for network analyses (Figure 6). The following active interaction sources were used: experiments, databases, co-expression, neighborhood, and gene fusion. The minimum interaction score was set to 0.7 (high confidence). The clusters were then characterized by biological processes and cellular localization using Gene Ontology (GO) [59] and the central gene nodes in each cluster were selected. Central gene nodes were identified as those with the highest number of connections with other genes in the network, a method used to increase the likelihood of identifying biomarker candidates that are fundamental to biological processes [8].

**Figure 6.**
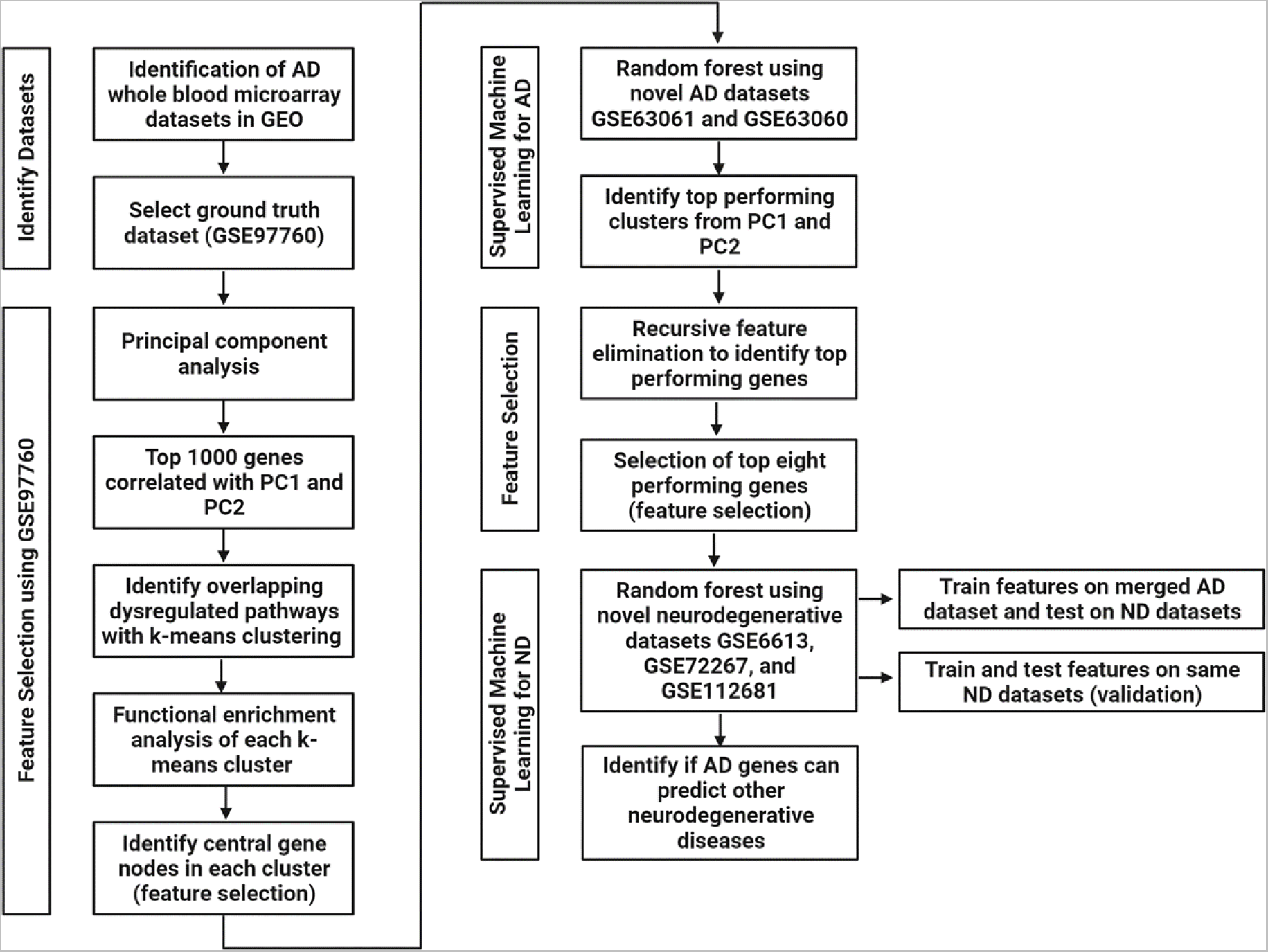
Flow chart of the methodological pipeline. Abbreviations: PC: principal component, ND: neurodegenerative diseases.

### 4.4. Supervised machine learning to identify the best predictors of AD

Once the central gene nodes from each k-means cluster were identified, we applied supervised machine learning (random forest) to determine which central gene nodes (features) were best able to distinguish AD patients from cognitively healthy controls. To do this, we used two novel AD datasets to train and test the models: GSE63061 and GSE63060. Both datasets, respectively, were split into training (70%) and testing (30%) datasets. The training datasets were used to perform model training, tuning, and validation. A 5-fold cross-validation repeated three times was used to improve model accuracy and identify the top performing gene biomarker predictors [60]. The final evaluation of our random forest models was done on withheld testing datasets. Performance indicators used to evaluate our models included sensitivity (correctly identifies AD patients) and precision (quality of positive AD prediction, i.e. number of AD patients / total number of predicted AD patients (true and false)). Using these metrics to determine the diagnostic utility of biomarker tests is particularly important because (a) it reduces the likelihood of producing a false negative outcome (someone who does have AD has been identified as being healthy) and (d) assesses the probability that a person with a positive result indeed has AD [61]. We did, however, also report additional performance metrics including AUC (ability to distinguish between AD patients and cognitively healthy controls) and specificity (correctly identified healthy controls). Random forest modelling was performed in R Studio v1.2.5033 using libraries rpart, caret, and pROC.

### 4.5. Feature selection to identify the genes that contribute to AD prediction

Within each of the best predictive models for the PC1 and PC2 clusters, respectively, we wanted to identify the genes that contributed the most to our models’ ability to predict AD cases. To do this, we performed RF-RFE and examined variable importance at the peak performance of each model (Figure 6). This method has previously been used to successfully identify important genes in transcriptomic data [62]. We then took the top eight important genes from each model to use as features in our neurodegenerative disease models (Figure 6).

### 4.6. Supervised machine learning to identify if AD gene biomarkers are unique to AD or generalizable to other neurodegenerative diseases

To identify whether our AD blood-based gene biomarkers were specific to AD, we tested whether the top eight important genes from each of the PC1 and PC2 clusters, respectively, were predictive of PD and ALS. We first identified three additional datasets in the GEO database that used microarray to analyze whole blood from patients with PD (GSE6613 and GSE72267) and ALS (GSE112681). Importantly, we did identify datasets examining other dementias, including behavioral variant frontotemporal dementia, and there was a substantial class imbalance whereby the number of healthy controls significantly outweighed the number of patients. Our modelling methods are not possible with such imbalanced datasets, and these were therefore excluded from our analyses. These datasets were processed in the same way as the AD datasets described above in section 4.2.

We then used the eight top important genes from PC1 and PC2 clusters in AD in a random forest model (Figure 6). We first merged the two AD datasets (GSE63061 and GSE63060) into a single dataset using z-scores as we’ve done previously [8]. This merged dataset was then used as a training dataset to train, tune, and validate our models. A 5-fold cross-validation repeated three times was also used. We then used each of the three neurodegenerative disease datasets, respectively, to test these AD-trained models (Figure 6). We also did a second validation experiment where our random forest models were both trained and tested on each of the neurodegenerative disease datasets (Figure 6). Each dataset was split into a 70% training, fine tuning, and validation dataset and a 30% withheld dataset for testing the models. As before, performance indicators used to evaluate our models included sensitivity (correctly identifies PD or ALS patients) and precision (quality of positive PD or ALS prediction, i.e. number of PD or ALS patients / total number of predicted PD or ALS patients (true and false)).

## Author Contributions

Conceptualization: A.S. & C.A.F.; methodology: A.S. & C.A.F.; data curation: C.A.F. software: A.S.; formal analysis: A.S., & C.A.F.; investigation: A.S., S.T., J.S., A.C., H.M.W., S.J.A., F.D., T.A.C.,J.K.I., F.I., S.K., E.D., C.D-S., S.R., N.M.R., D.C., W.A.G., R.H.S., D.A.B., & C.A.F.; resources: C.A.F., writing – original draft preparation: A.S. & C.A.F., writing – review and editing: A.S., S.T., J.S., A.C., H.M.W., S.J.A., F.D., T.A.C., J.K.I., F.I., S.K., E.D., C.D-S., S.R., N.M.R., D.C., W.A.G., R.H.S., D.A.B., & C.A.F.; supervision: C.A.F.; project administration: C.A.F.

## Data Availability Statement

All data in this study is publicly accessible in the Gene Expression Omnibus (GEO) database under accession numbers: GSE97760, GSE63061, GSE63060, GSE6613, GSE72267, and GSE112681. All data generated from our analyses is available in the Supplementary Tables. Code is available on A.S.’s Github at https://github.com/Art83.

