## Supplementary Figures for "Blood-based transcriptomic biomarkers are predictive of neurodegeneration rather than Alzheimer’s disease"

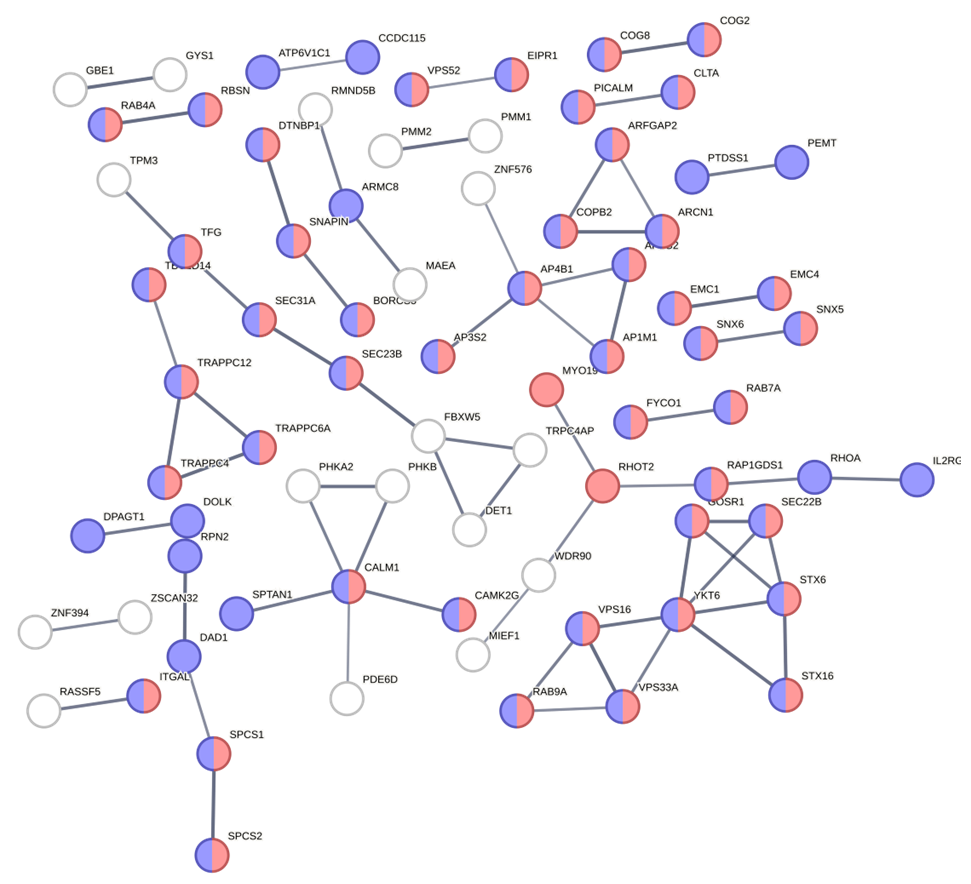

**Supplementary Figure 1.** Characterization of the biological function and cellular localization of component 1 (PC1) k-means cluster 1 (red) using Gene Ontology and STRING. The STRING network analysis includes the following interaction sources: experiments, databases, co-expression, neighborhood, and gene fusion. The minimum interaction score was set to 0.7 (high confidence) and disconnected nodes in the network are hidden. Thickness of the line indicates the confidence in the interaction. The red nodes indicate genes involved in cellular localization (FDR = 3.61e-11) and the blue nodes indicate cellular localization of endomembrane system (FDR = 6.02e-22).

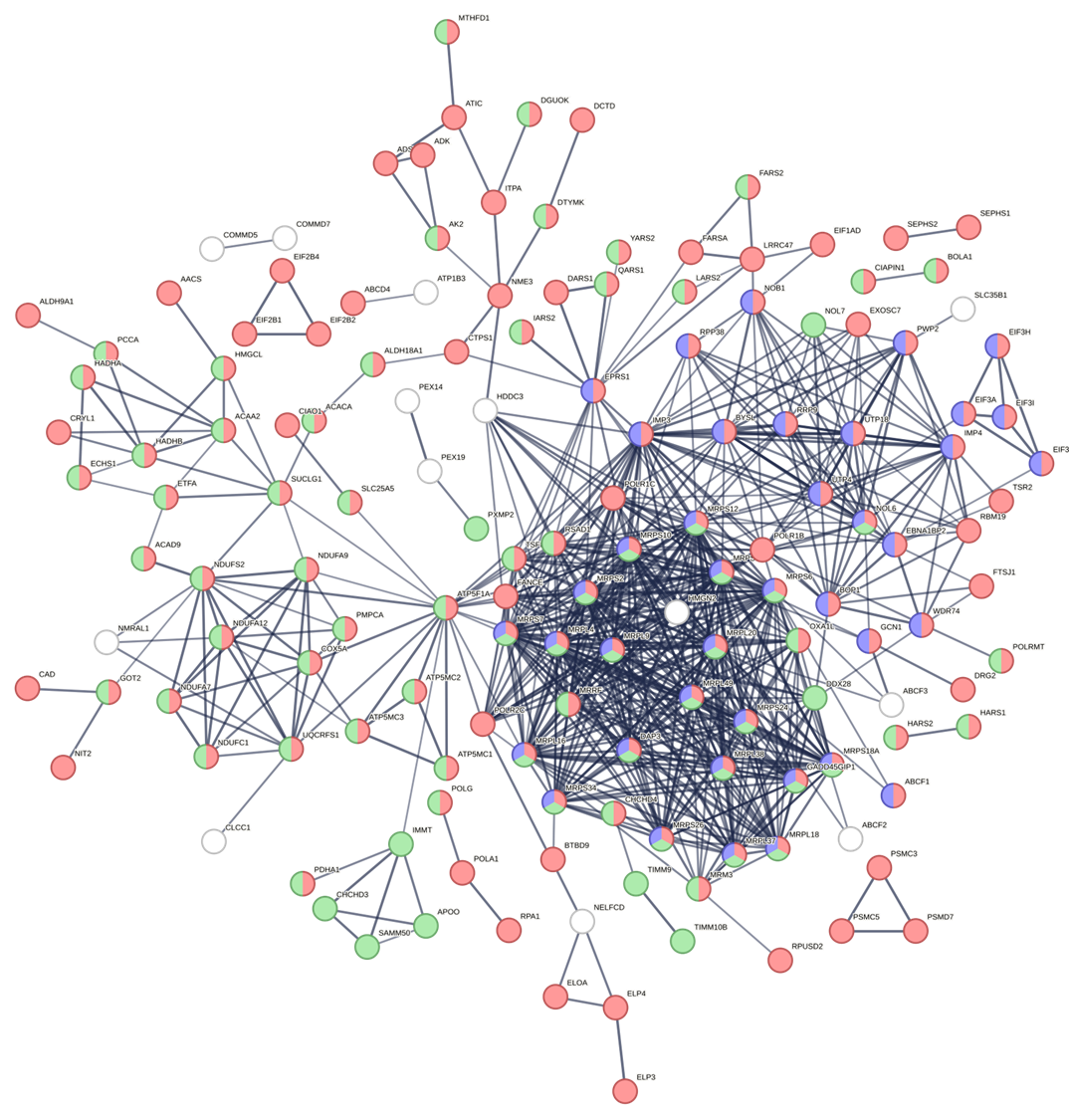

**Supplementary Figure 2.** Characterization of the biological function and cellular localization of component 1 (PC1) k-means cluster 2 (yellow) using Gene Ontology and STRING. The STRING network analysis includes the following interaction sources: experiments, databases, co-expression, neighborhood, and gene fusion. The minimum interaction score was set to 0.7 (high confidence) and disconnected nodes in the network are hidden. Thickness of the line indicates the confidence in the interaction. The red nodes indicate genes involved in metabolic processes (FDR = 5.29e-25), the green nodes indicate cellular localization of mitochondrion (FDR = 2.87e-40), and the blue nodes indicate ribonucleoprotein complex (FDR = 5.93e-12).

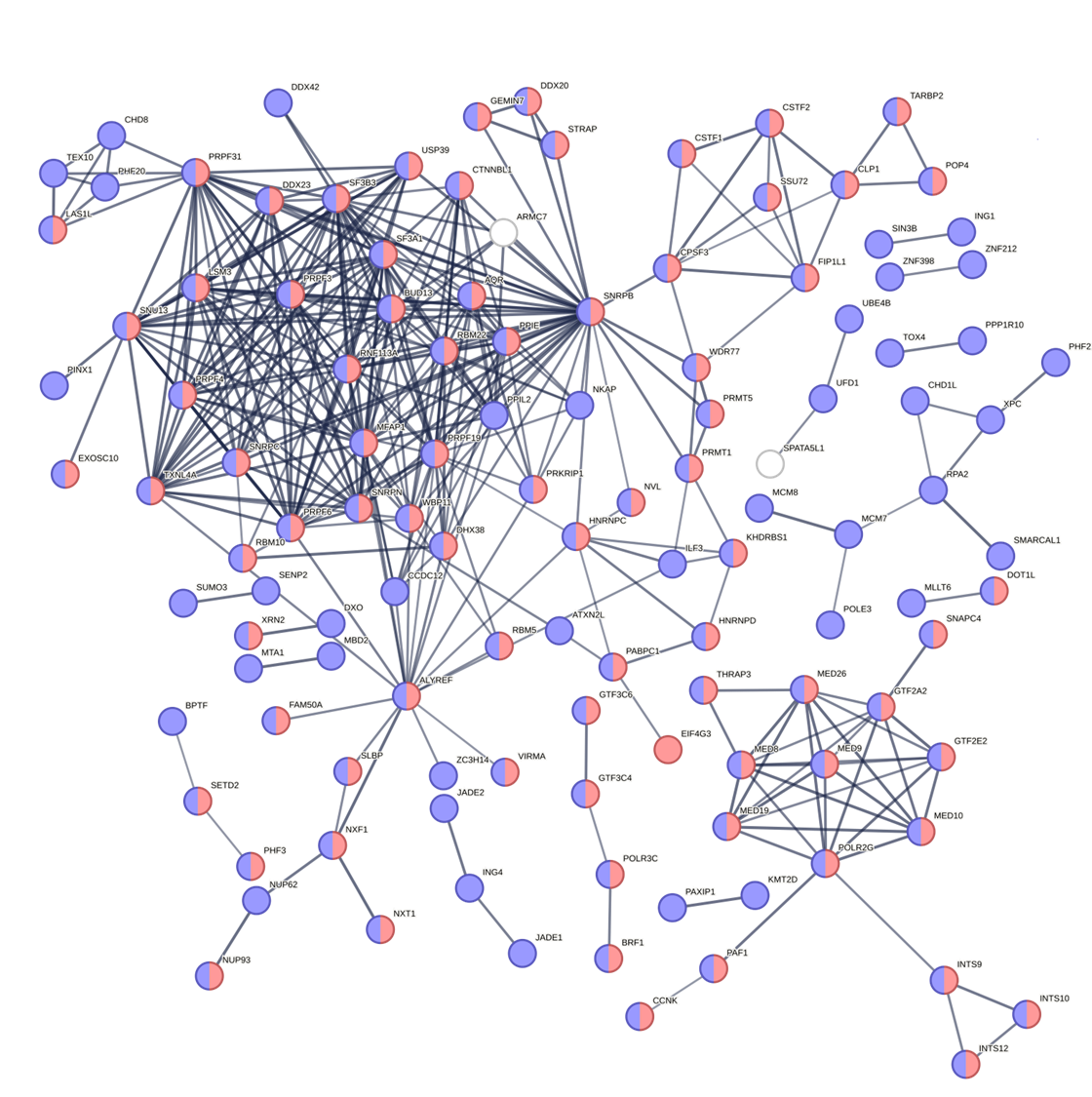

**Supplementary Figure 3.** Characterization of the biological function and cellular localization of component 1 (PC1) k-means cluster 3 (green) using Gene Ontology and STRING. The STRING network analysis includes the following interaction sources: experiments, databases, co-expression, neighborhood, and gene fusion. The minimum interaction score was set to 0.7 (high confidence) and disconnected nodes in the network are hidden. Thickness of the line indicates the confidence in the interaction. The red nodes indicate genes involved in gene expression (FDR = 3.87e-28), the blue nodes indicate cellular localization of the nucleus (FDR = 1.75e-44).

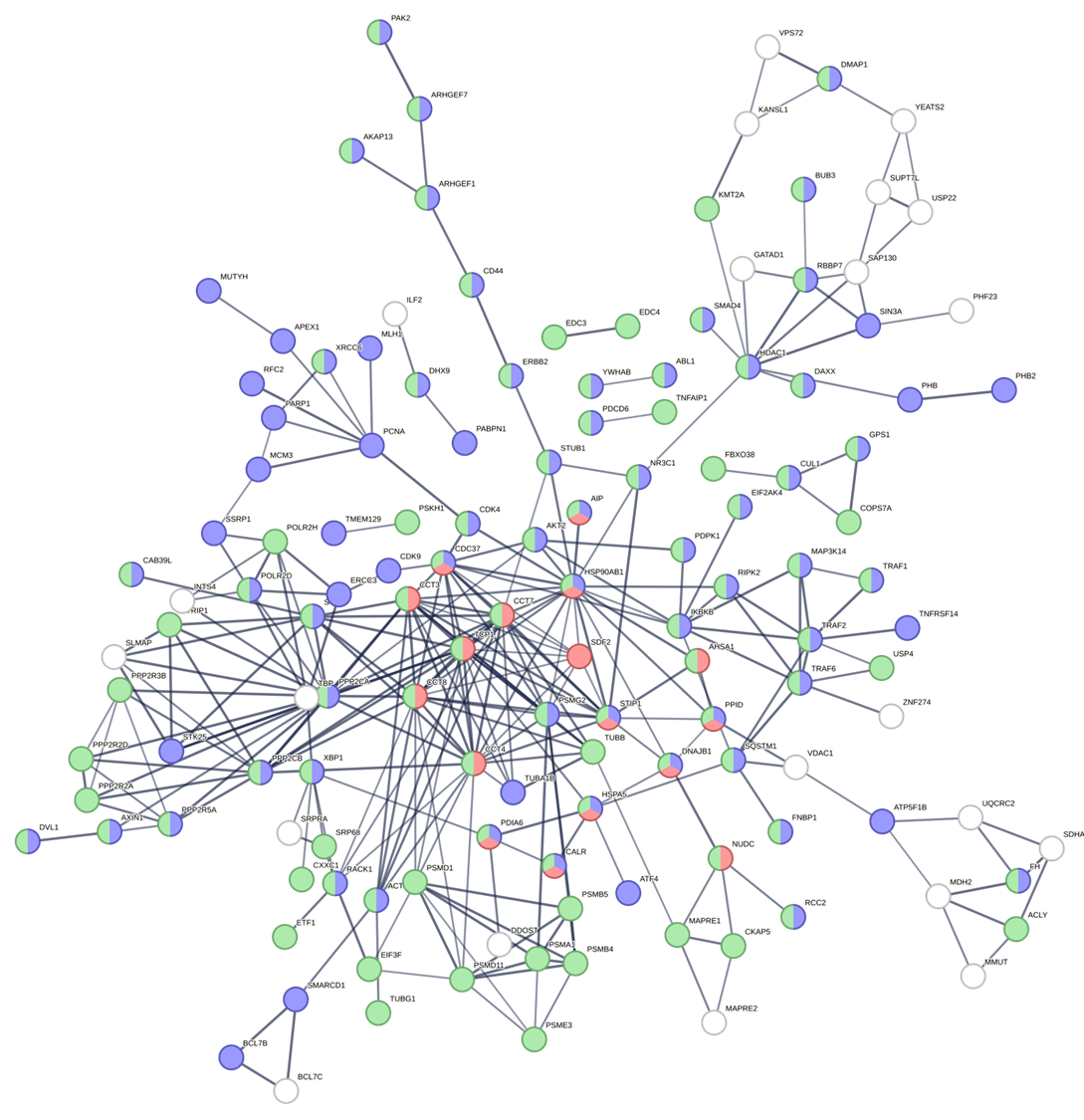

**Supplementary Figure 4.** Characterization of the biological function and cellular localization of component 1 (PC1) k-means cluster 4 (blue) using Gene Ontology and STRING. The STRING network analysis includes the following interaction sources: experiments, databases, co-expression, neighborhood, and gene fusion. The minimum interaction score was set to 0.7 (high confidence) and disconnected nodes in the network are hidden. Thickness of the line indicates the confidence in the interaction. The red nodes indicate genes involved in protein folding (FDR = 2.80e-9), the blue nodes indicate genes involved in cellular response to stimulus (FDR = 1.69e-9), and the green nodes indicate cellular localization of cytosol (FDR = 1.19e-16).

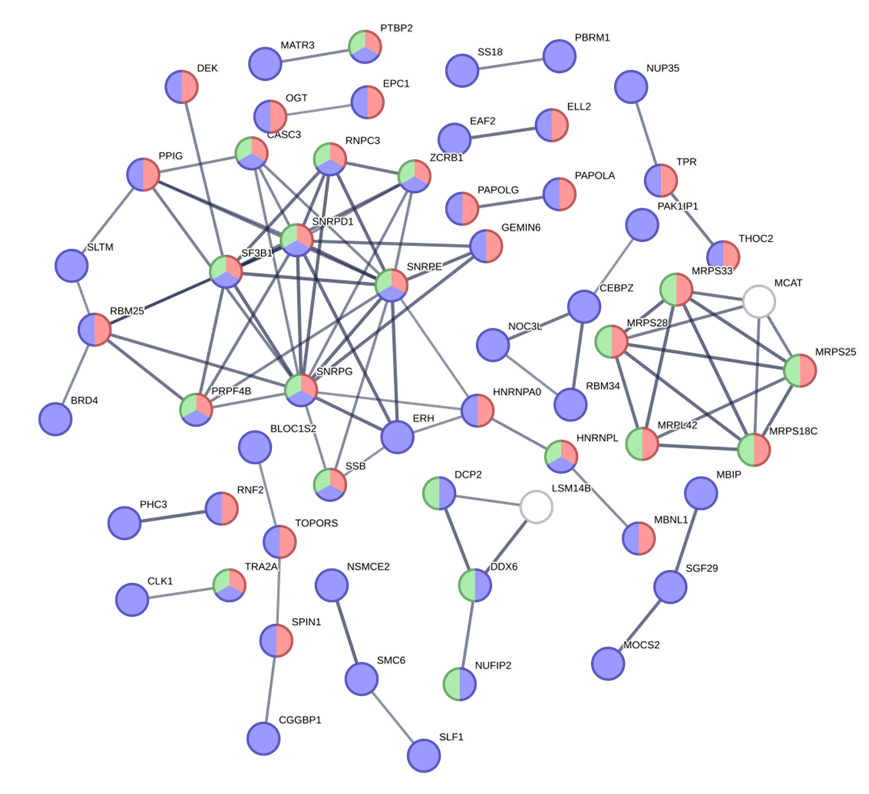

**Supplementary Figure 5.** Characterization of the biological function and cellular localization of component 1 (PC2) k-means cluster 1 (red) using Gene Ontology and STRING. The STRING network analysis includes the following interaction sources: experiments, databases, co-expression, neighborhood, and gene fusion. The minimum interaction score was set to 0.7 (high confidence) and disconnected nodes in the network are hidden. Thickness of the line indicates the confidence in the interaction. The red nodes indicate genes involved in gene expression (FDR = 2.32e-9), the blue nodes indicate cellular localization to the nucleus (FDR = 1.83e-21), and the green nodes indicate cellular localization of ribonucleoprotein complex (FDR = 0.00024).

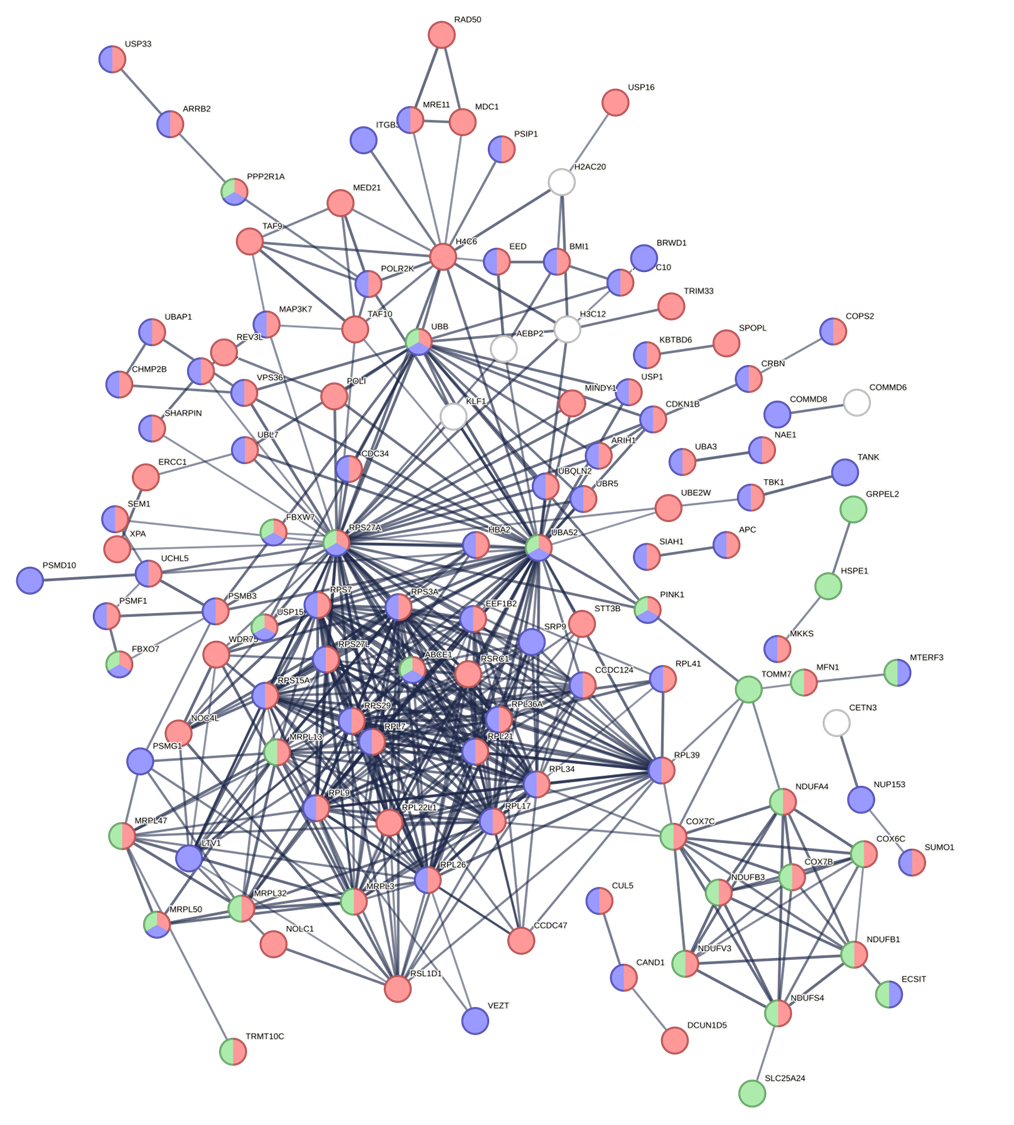

**Supplementary Figure 6.** Characterization of the biological function and cellular localization of component 1 (PC2) k-means cluster 2 (yellow) using Gene Ontology and STRING. The STRING network analysis includes the following interaction sources: experiments, databases, co-expression, neighborhood, and gene fusion. The minimum interaction score was set to 0.7 (high confidence) and disconnected nodes in the network are hidden. Thickness of the line indicates the confidence in the interaction. The red nodes indicate genes involved in metabolic process (FDR = 2.58e-22), the blue nodes indicate cellular localization to the cytosol (FDR = 1.78e-16), and the green nodes indicate cellular localization of mitochondrion (FDR = 4.40e-6).

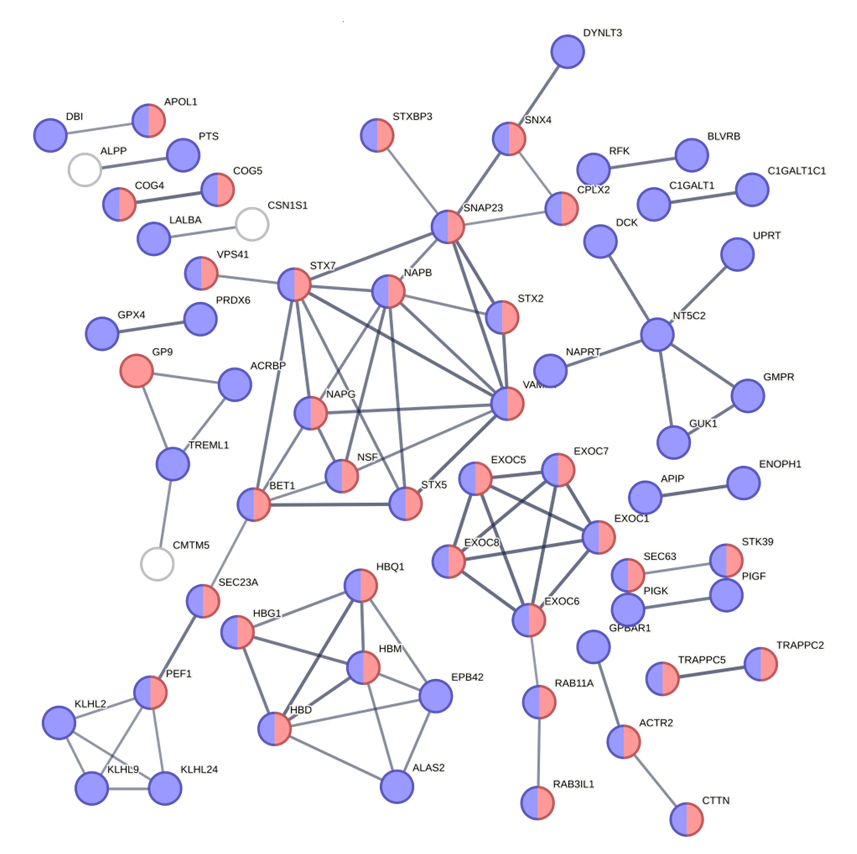

**Supplementary Figure 7.** Characterization of the biological function and cellular localization of component 1 (PC2) k-means cluster 3 (green) using Gene Ontology and STRING. The STRING network analysis includes the following interaction sources: experiments, databases, co-expression, neighborhood, and gene fusion. The minimum interaction score was set to 0.7 (high confidence) and disconnected nodes in the network are hidden. Thickness of the line indicates the confidence in the interaction. The red nodes indicate genes involved in transport (FDR = 4.58e-6) and the blue nodes indicate cellular localization to the cytoplasm (FDR = 4.19e-10).

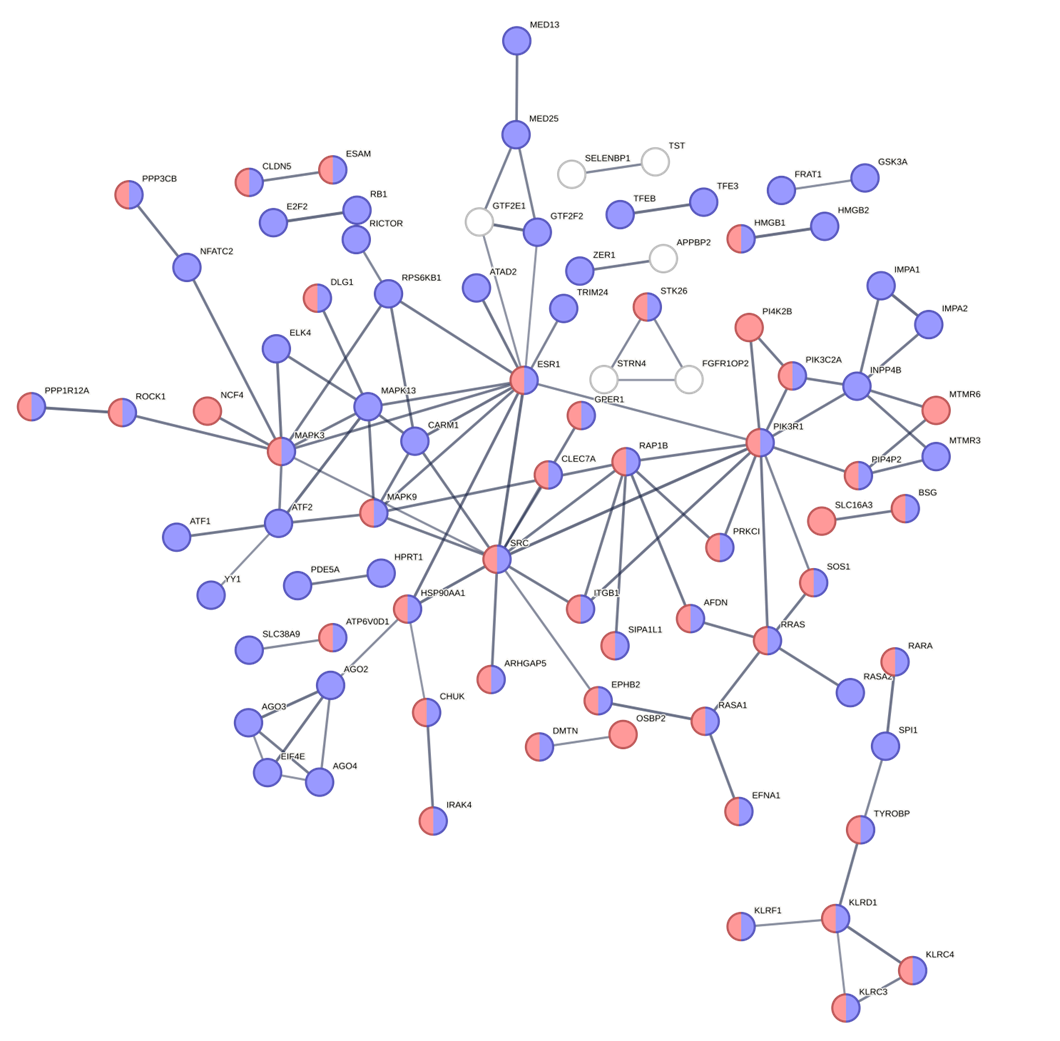

**Supplementary Figure 8.** Characterization of the biological function and cellular localization of component 1 (PC2) k-means cluster 4 (blue) using Gene Ontology and STRING. The STRING network analysis includes the following interaction sources: experiments, databases, co-expression, neighborhood, and gene fusion. The minimum interaction score was set to 0.7 (high confidence) and disconnected nodes in the network are hidden. Thickness of the line indicates the confidence in the interaction. The red nodes indicate genes involved in regulation of cellular process (FDR = 2.49e-10) and the blue nodes indicate cellular localization to the plasma membrane (FDR = 0.00062).
